# Exploring barriers and facilitators to physical activity during the COVID-19 pandemic: a qualitative study

**DOI:** 10.1101/2022.05.18.22275240

**Authors:** Charlotte Roche, Abigail Fisher, Daisy Fancourt, Alexandra Burton

## Abstract

**Objectives:** Quantitative data show that physical activity (PA) reduced during the COVID-19 pandemic, with differential impacts across demographic groups. Qualitative research is limited, so reasons for this have not been explored in-depth. This study aimed to understand barriers and facilitators to PA during the pandemic, focusing on groups more likely to have been affected by restrictions, and to map these onto the Capability, Opportunity, Motivation Model of Behaviour (COM-B).

**Design:** Semi-structured qualitative interview study.

**Methods:** One-to-one telephone/videocall interviews were conducted with younger (aged 18-24) and older adults (aged 70+), those with long-term physical health conditions or mental health conditions, and parents of young children, probing about their experiences of PA. Barriers and facilitators were identified using reflexive thematic analysis, and themes were mapped onto COM-B dimensions.

**Results:** 116 participants were included (18-93 years old, 61% female, 71% White British). Key themes were the importance of the outdoor environment, impact of COVID-19 restrictions, fear of contracting COVID-19, and level of engagement with home exercise. Caring responsibilities and conflicting priorities were a barrier. PA as a method of socialising, establishing new routines, and the importance of PA for protecting mental health were motivators. Most themes mapped onto the physical opportunity (environmental factors) and reflective motivation (evaluations and plans) COM-B domains.

**Conclusions:** Future interventions should increase physical opportunity and reflective motivation for PA during pandemics, to avoid further negative health outcomes following periods of lockdown. Strategies could include tailoring PA guidance depending on location and giving education on the health benefits of PA.

**Statement of Contribution:** *What is already known on this subject?:*

- Physical activity (PA) levels reduced during the COVID-19 pandemic.
- The extent of this reduction varied across demographic groups.
- Very few qualitative studies have explored reasons for these changes.

*What does this study add?:*

- Novel interview data, giving context to existing quantitative data.
- Insight into which themes were important for different demographic groups.
- Suggestions for increasing PA in future pandemics, by mapping findings to a theoretical framework.

## Introduction

The World Health Organisation (WHO) declared COVID-19 a global pandemic in March 2020 (WHO, 2021). In response, the UK government introduced measures to reduce transmission, including social distancing, closure of exercise and hospitality venues, and full lockdowns where the population was advised to ‘stay at home’. Under the strictest guidance, physical activity (PA) outside of the home was limited to once per day within the local area (GOV UK, 2021).

Many quantitative studies have examined the effect of pandemic restrictions on PA. A systematic review of 66 observational studies found that PA reduced during periods of lockdown (Stockwell et al., 2020). However, there are indicators of differential (or conflicting) findings when studies have explored effects by demographic groups. For example, a longitudinal survey and smartphone tracking study found substantial declines in PA for younger adults (Savage et al., 2021; McCarthy, Potts & Fisher, 2021). But whilst many studies suggested that older adults may have been more likely to maintain, or even increase, their PA during the pandemic (Smith et al., 2020; Rogers et al., 2020; To et al., 2021; McCarthy et al., 2021), others showed a stronger decline in older adults (Wang et al., 2020; Schmidt & Pawlowski, 2021; Strain et al., 2022).

The pandemic restrictions were more profoundly experienced for some demographic groups, likely resulting in greater reductions in PA. For example, over 2.2 million individuals living with long-term health conditions (LTCs) in the UK were advised to ‘shield’ (not leave their home or garden at all) (Office for National Statistics, 2020). Observational studies suggest corresponding declines in PA for those with LTCs (Assaloni et al., 2020; Ghosh, Arora, Gupta, Anoop & Misra, 2020; Roberts-Lewis, Ashworth, White & Rose, 2021). Similarly, cross-sectional studies suggest that individuals with mental health conditions (MHCs) showed lower levels of PA during the pandemic (Carvalho, Borges-Machado, Pizarro, Bohn & Barros, 2021; Savage et al., 2021; Mishra et al., 2021; Creese et al., 2021). Further, parents of younger children faced additional caring responsibilities due to closures of childcare settings and schools (Institute for Government, 2021), which may have led to them being less active (Mata et al., 2021; Robbins & Ling, 2022; Curtis et al., 2021).

Despite the large body of quantitative data, very few qualitative studies have explored the impact of the pandemic on PA, and most have focused specifically on older adults. One study explored the experiences of 17 older adults living in Australian retirement villages. Barriers to PA included closure of exercise facilities, cancellation of group activities and lack of equipment; while facilitators were wanting to maintain fitness, enjoyment, experiencing positive emotions after PA, exercising with someone, access to technology and provision of exercise routines (Ng et al., 2021). Interviews conducted with six general population older adults in France (Goethals et al., 2020) and 25 in the US (Greenwood-Hickman et al., 2021) found that they stopped attending exercise classes due to fear of COVID-19 infection. Additional barriers included concerns about safety, working from home and not having knowledge of/interest in online PA resources (Greenwood-Hickman et al., 2021; Goethals et al., 2020). A UK study used a mixed-methods approach with telephone interviews to understand reasons for pandemic-related change in PA levels for 26 adults with LTCs. Reasons included having to ‘shield’, fear of catching COVID-19, lack of garden access and closure of exercise facilities (Smith et al., 2021). Free text coding of survey data from 1214 Irish adolescents found that facilitators for PA during the pandemic included not having to attend school, having little else to do and prioritising health; while barriers included low motivation, lack of routine, cancelled club activities and health concerns (Ng, Cooper, McHale, Clifford & Woods, 2020). However, data were gathered using online surveys which, while valuable, limits the depth of information that participants can provide.

Furthermore, application of qualitative findings to theoretical frameworks is limited. The capability, opportunity and motivation model of behaviour (COM-B) is particularly useful, as barriers and facilitating factors map directly onto intervention functions, which is helpful for informing intervention development for future pandemics and health emergencies. The model suggests that capability (knowledge and skills), opportunity (social and environmental factors) and motivation (cognitive processes directing behaviour) are key for engagement inb a target behaviour (Michie, van Stralen & West, 2011). The COM-B constructs of capability and motivation quantitatively explained a large proportion of variance in moderate to vigorous PA prior to the pandemic (Howlett, Schulz, Trivedi, Troop & Chater, 2019), and the COM-B model was successfully utilised in the COVID-19 Social Study (CSS) to help understand the psychological factors influencing compliance with social distancing guidance (Burton et al., 2022). One systematic review explored correlates of PA and sedentary behaviour during the pandemic and mapped the themes to the COM-B model (Knight et al., 2021). This study identified a range of individual psychological factors (e.g., motivation, mental health), social level factors (e.g., social support) and environmental factors (e.g., area of residence) as key for PA and sedentary behaviour. However, the review was largely based on quantitative data, which does not give the depth of perspectives accessible through qualitative methods. Additionally, while authors suggested that changes to all domains of the COM-B model are important for facilitating PA engagement, particularly physical opportunity and psychological capability, they did not make specific recommendations for how this could be done.

The aim of this study was therefore to qualitatively explore barriers and facilitators to PA during the COVID-19 pandemic across different demographic groups whose participation in PA may have been particularly affected, and to map these onto the COM-B model.

## Methods

### Design

This was part of the qualitative arm of the UCL-COVID-19 Social Study (CSS) (COVID-19 Social Study, 2020). Although the overarching aim of the CSS was to explore the impact of COVID-19 restrictions on mental health and wellbeing, a planned sub-aim was to explore the impact on PA.

### Participants

Participants were recruited using convenience sampling by advertising the study on the CSS website, newsletter, and social media, through personal contacts and via third party organisations through the MARCH Mental Health Research network (MARCH Network, 2022). The target groups for recruitment in the qualitative arm of the CSS were those whose mental health and wellbeing was hypothesised to have been particularly impacted by the pandemic and associated restrictions (e.g., adolescents, younger adults, older adults, those with LTCs, parents of young children, those with MHCs, frontline healthcare workers, non-health key workers, those in the gig economy and adults experiencing long-term effects of COVID-19). Participants were eligible if they were living in the UK, identified with one or more of the target groups, had good understanding of English and were willing and able to provide informed consent (or parental consent was given for adolescents). Participant response rates were not recorded.

Between May 2020 and January 2021, over 200 participants in the UK were interviewed. The COVID-19 restrictions that were in place during the data collection period are described in Table 1. For the current study, interviews were included from groups who we considered (based on existing quantitative data) were likely to be especially impacted by the pandemic in terms of their PA and to have discussed it in enough depth to code (e.g., interview data with frontline workers and those in the gig economy related more to the effects of the pandemic on work, rather than PA). Groups we focused on included young adults aged 18-24 (Savage et al., 2021; McCarthy et al., 2021), older adults over the age of 70 (Wang et al., 2020; Schmidt & Pawlowski, 2021), those with LTCs (Office for National Statistics, 2020; Assaloni et al., 2020; Ghosh et al., 2020; Roberts-Lewis et al., 2021) or MHCs (Carvalho et al., 2021; Savage et al., 2021; Mishra et al., 2021; Creese et al., 2021) and parents of young children (Institute for Government, 2021; Mata et al., 2021; Robbins & Ling, 2022; Curtis et al., 2021). Ethical approval was provided by the University College London (UCL) research ethics committee ((project identifier 14895/005), and all participants provided informed written consent. Participants were offered a £20 voucher for participation.

**Table 1.** Timeline of COVID-19 restrictions in England 2020-2021. Restrictions were similar across all countries within the UK, but dates and precise details may vary.

| **Date** | **Timeline of COVID-19 News and Restrictions** |
| --- | --- |
| 21 Mar 2020 | Shielding for the clinically extremely vulnerable and over 70s is introduced and is predicted to last for 12 weeks. |
| 23 Mar 2020 | National lockdown begins. Legal exemptions for leaving home include essential shopping, going to work, medical reasons, exercising once per day. |
| 10 May 2020 | Those who cannot work from home encouraged to return to work but avoid public transport.<br>Unlimited outdoor exercise is permitted. |
| 13 May 2020 | Outdoor recreation added as a legal exemption or leaving home.<br>People allowed to meet someone from one other household outside. |
| 1 Jun 2020 | 'Stay at home' order removed.<br>Outdoor gatherings of six people from different households permitted.<br>Indoor gatherings of two people from different households permitted.<br>Schools begin to reopen gradually. |
| 23 Jun 2020 | Announcement that hospitality, some leisure facilities and attractions can reopen.<br>Weddings and domestic holidays allowed. |
| 4 Jul 2020 | Local lockdowns are introduced for areas where the rates of positive cases of COVID-19 are higher. |
| 14 Sep 2020 | Socialising is limited to groups of six people. Gatherings of above six people are banned. |

| Date | Timeline of COVID-19 News and Restrictions |
| --- | --- |
| 22 Sep 2020 | Return to working from home.<br>Hospitality venues must close at 10pm. |
| 14 Oct 2020 | Tier system is introduced where different regions of the country will live under different restrictions depending on local infection rates.<br>• Tier 1 (medium alert) – rule of six when socialising, work from home if possible.<br>• Tier 2 (high alert) – rule of six outside but no inside contact with other households, work from home if possible.<br>• Tier 3 (very high alert) – socialising with other households not permitted in most settings, work from home if possible, sports facilities are open but close contact sports should be avoided, indoor exercise only permitted with your household. |
| 5 Nov 2020 | Second national lockdown begins. |
| 2 Dec 2020 | End to national lockdown and return to tier system. |
| 20 Dec 2020 | Tier 4 introduced – restrictions are similar to a lockdown with a ‘stay at home’ order, non-essential shops, hospitality and sports facilities are closed. |
| 6 Jan 2021 | Third national lockdown begins. |
| 8 Mar 2021 | Schools begin to reopen.<br>Two people are permitted to meet for outdoor recreation. |
| 29 Mar 2021 | Groups of six people or two households can meet outside.<br>Outdoor sports facilities reopen. |
| 12 Apr 2021 | Non-essential shops, outdoor hospitality and gyms re-open. |
| 17 May 2021 | Groups of six people or two households can meet inside.<br>Most indoor venues reopen. |
| 19 Jul 2021 | Most COVID-19 restrictions end.<br>Social distancing guidance and self-isolation rules remain. |
| 27 Nov 2021 | First cases of new ‘Omicron’ COVID-19 variant detected in the UK, which is expected to be more transmissible than previous variants. |
| 8 Dec 2021 | COVID-19 restrictions re-introduced in the UK as part of winter ‘Plan B’. These include; working from home where possible, proof of vaccination or negative lateral flow test required for large gatherings and mandatory use of face coverings in most public indoor settings. |

*Note.* As outlined in House of Commons Library (2021). Coronavirus: A history of English lockdown laws. Retrieved from https://researchbriefings.files.parliament.uk/documents/CBP-9068/CBP-9068.pdf

### Qualitative Interviews

Interviews were conducted one-to-one via video or telephone call by a member of the CSS team with no prior relationship with any participant they interviewed. Interviewers were male and female postgraduate researchers who had prior experience of conducting qualitative interviews with people experiencing physical and mental health difficulties, and vulnerable groups. Interviews followed a topic guide designed to explore the impact of the pandemic on participants’ mental health, social lives and worries for the future, the findings of which have been published (Burton, McKinlay, Aughterson & Fancourt, 2021; McKinlay, May, Dawes, Fancourt & Burton, 2021; McKinlay, Fancourt & Burton, 2021; Fisher, Roberts, McKinlay, Fancourt & Burton, 2021; Dawes, May, McKinlay, Fancourt & Burton, 2021; Burton, McKinlay, Aughterson & Fancourt, 2021). Questions also prompted participants to consider the effect of the pandemic on their PA (prompts are shown in Figure 1). No repeat interviews were conducted, and no field notes were made during the interviews.

**Figure 1.**
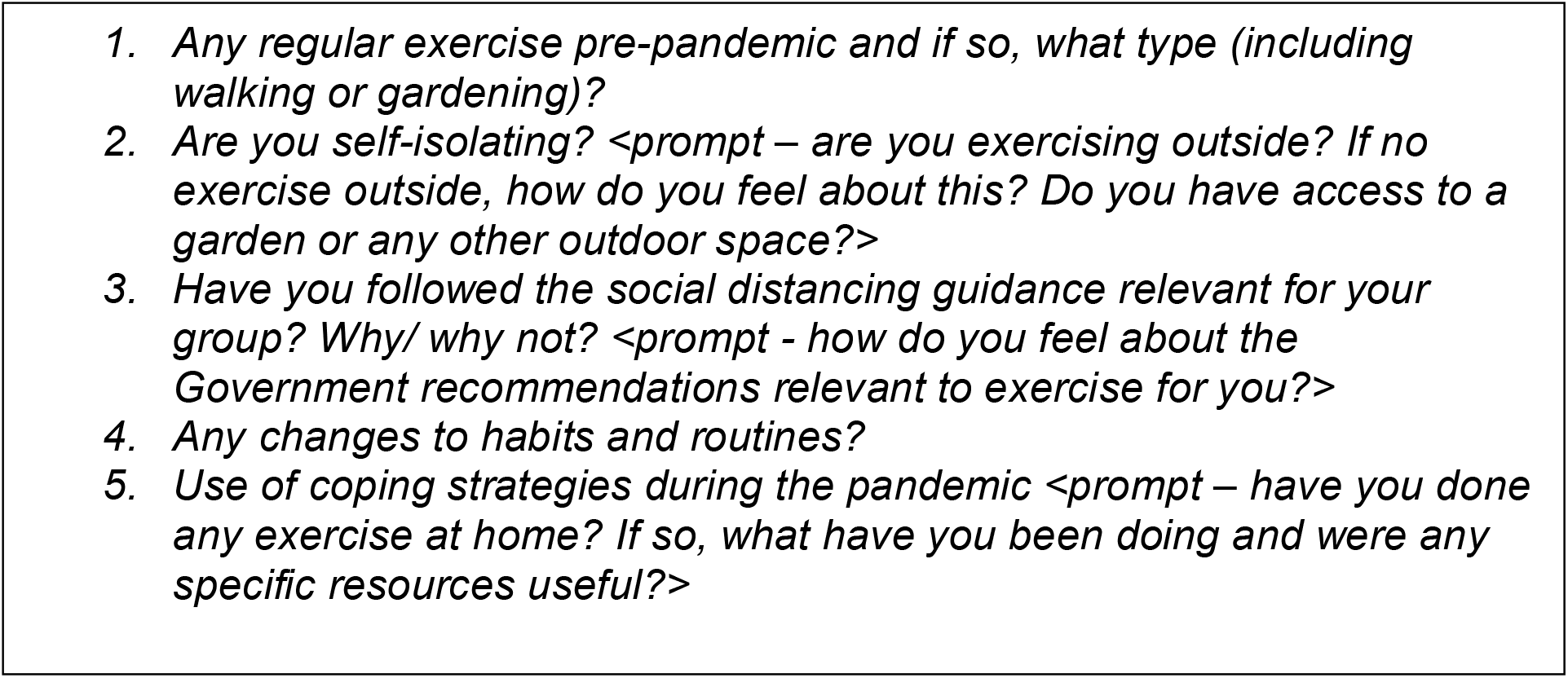
Prompts from the CSS topic guide relating to pandemic-related changes in PA.

Interviews were audio recorded and transcribed verbatim by an external service with a UCL data sharing agreement. Transcripts were de-identified prior to analysis.

### Analysis

Transcripts were analysed using reflexive thematic analysis (Braun & Clarke, 2019) with a phenomenological approach, aiming to use lived experiences to evoke meaning and identification of themes (Sundler, Lindberg, Nilsson & Palmér, 2019). Thematic analysis was selected due to its flexible nature which allows for inductive data-driven findings. Initially, transcripts were read by CR for familiarisation. Then, initial codes were generated inductively from a randomly selected sub-set of six transcripts, second coded by AB and AF. Coding was carried out independently before the researchers met to discuss initial codes and findings, and the concepts identified were generally consistent. Therefore, coding and the subsequent processes of thematic analysis; generation, reviewing and grouping of themes (Braun & Clarke, 2006; Braun & Clarke, 2021) were conducted for the remaining interviews by CR using Microsoft Excel and NVIVO12. NVIVO12 was used to code the qualitative interviews and to group the codes into broader themes. Microsoft Excel was then used to organise the themes into barriers, facilitators or both depending on participant circumstance. Themes were also grouped by their relation to physical or psychological capability, social or physical opportunity and automatic or reflective motivation (Michie et al., 2011). The theoretical domains framework was used to support decision making on these COM-B groupings; a group of 14 theoretical domains which are linked to the COM-B model (Atkins et al., 2017). Upon finalisation of themes, CR reflected on which themes appeared important or not important within and across the different demographic groups.

The research team met regularly during the data analysis period. There were no significant disagreements between researchers throughout. Participants were not involved with the design, analysis, interpretation, or dissemination stages of this study.

## Results

### Descriptive results

116 participants were included in the study. 71 (61%) were female, and the mean age was 50 years (SD = 20). See Table 2 for a breakdown by demographic group. Participants predominantly identified as ‘White British’ (71%), although a range of ethnicities were represented. Most participants were educated to degree level (73%). Mean interview length was 56 minutes (Range= 14 – 149 minutes).

**Table 2.** Participant demographics for the current study.

|  | Young Adult | Older Adult | MHC | LTC | Parents of Young Children | Total |
| --- | --- | --- | --- | --- | --- | --- |
| **Sample Size** | 16 | 20 | 22 | 33 | 25 | 116 |
| **Gender** |  |  |  |  |  |  |
| Male | 6 (37.50%) | 11 (55%) | 9 (40.91%) | 12 (36.36%) | 7 (28%) | 45 (38.79%) |
| Female | 10 (62.50%) | 9 (45%) | 13 (59.09%) | 21 (63.64%) | 18 (72%) | 71 (61.21%) |
| **Age (years)** |  |  |  |  |  |  |
| Mean | 21.06 | 79.30 | 43.23 | 56.36 | 40.32 | 49.50 |
| SD | 2.05 | 5.50 | 14.00 | 12.73 | 6.49 | 20.00 |
| **Ethnicity** |  |  |  |  |  |  |
| White British | 12 (75%) | 16 (80%) | 14 (63.64%) | 24 (72.73%) | 16 (64%) | 82 (70.70%) |
| White Irish | 0 | 0 | 1 (4.55%) | 0 | 2 (8%) | 3 (2.59%) |
| White & Asian | 0 | 0 | 2 (9.09%) | 1 (3.03%) | 1 (4%) | 4 (3.45%) |
| White & Black Caribbean | 1 (6.25%) | 0 | 0 | 1 (3.03%) | 0 | 2 (1.72%) |
| Black British Caribbean | 0 | 0 | 0 | 2 (6.06%) | 0 | 2 (1.72%) |
| Black British African | 0 | 0 | 2 (9.09%) | 0 | 1 (4%) | 3 (2.59%) |
| Indian | 1 (6.25%) | 0 | 2 (9.09%) | 2 (6.06%) | 2 (8%) | 7 (6.03%) |
| Chinese | 0 | 0 | 0 | 0 | 1 (4%) | 1 (0.86%) |
| Pakistani | 0 | 0 | 1 (4.55%) | 0 | 0 | 1 (0.86%) |
| Other | 2 (12.50%) | 4 (20%) | 0 | 3 (9.09%) | 2 (8%) | 11 (9.48%) |
| **Education** |  |  |  |  |  |  |
| No qualifications | 1 (6.25%) | 0 | 0 | 0 | 0 | 1 (0.86%) |
| Post-16 vocational course | 0 | 0 | 2 (9.09%) | 0 | 0 | 2 (1.72%) |
| GCSE/ equivalent | 0 | 3 (15%) | 1 (4.55%) | 3 (9.09%) | 1 (4%) | 8 (6.90%) |
| A-levels/ equivalent | 9 (56.25%) | 1 (5%) | 5 (22.73%) | 5 (15.15%) | 0 | 20 (17.24%) |
| Undergraduate degree/ professional qualification | 5 (31.25%) | 7 (35%) | 10 (45.45%) | 12 (36.36%) | 9 (36%) | 43 (37.07%) |
| Postgraduate degree | 1 (6.25%) | 9 (45%) | 4 (18.18%) | 13 (39.39%) | 15 (60%) | 42 (36.21%) |

### Results

Four themes (with seven subthemes) were identified as either barriers or facilitators for PA depending on participant circumstances: importance of the outdoor environment, the impact of COVID-19 restrictions, fear of contracting COVID-19 and related complications and level of engagement with home exercise. Caring responsibilities and conflicting priorities were identified as a barrier. Finally, three facilitators/motivators included: PA as a method of socialising, establishing new routines and the importance of PA for protecting mental health.

The themes are visually mapped onto the COM-B model dimensions in Figure 2. Most themes were mapped onto the domains of physical opportunity and reflective motivation including importance of the outdoor environment, impact of COVID-19 restrictions, level of engagement with home exercise and caring responsibilities and conflicting priorities. Establishing new routines and the importance of PA for protecting mental health related to reflective motivation alone. Fear of COVID-19 was mapped onto reflective and automatic motivation. Finally, PA as a method of socialising related to social opportunity. No themes related to physical or psychological capability.

**Figure 2.**
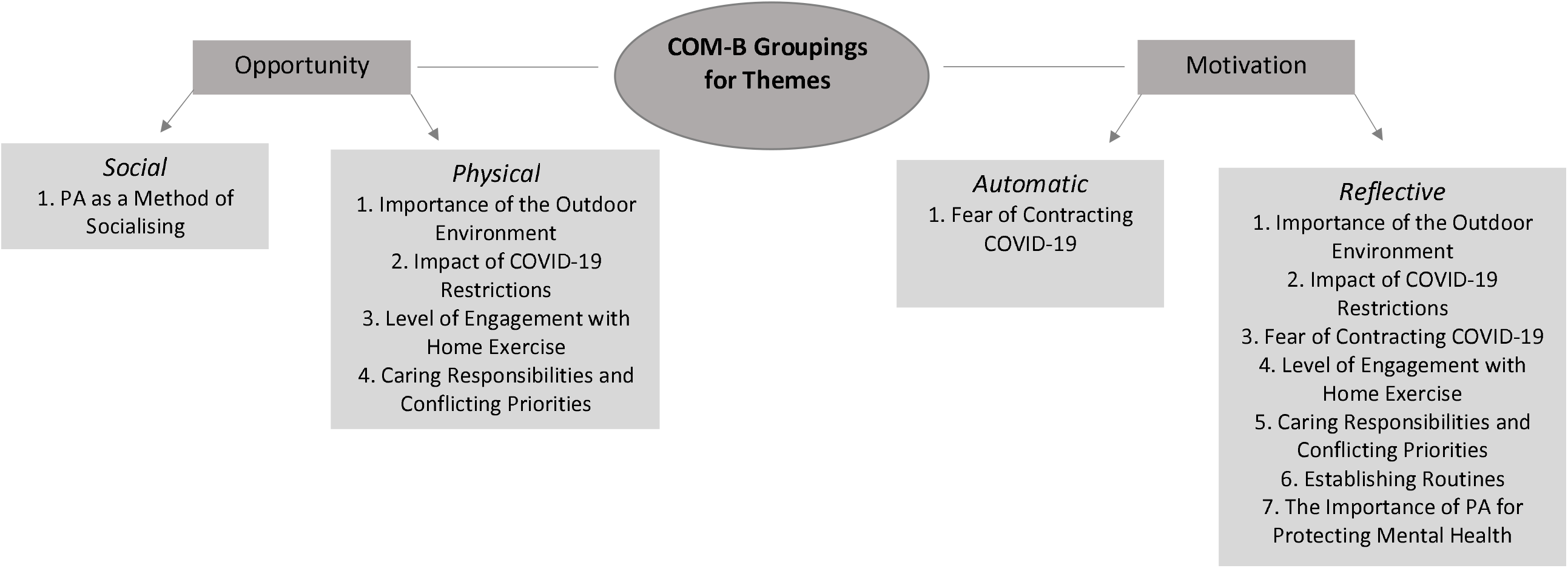
Themes mapped onto COM-B dimensions.

#### 1. Importance of the outdoor environment

The surrounding environment was key to many participants’ PA engagement and was one of the most important factors influencing PA. Garden access was a facilitator for exercise at home. Many participants described their local environment as a barrier or facilitator to PA depending on the surroundings and ability to socially distance. The weather was also raised as a key influence.

##### 1.1 Garden Access

For some participants, garden access provided an opportunity to engage in PA, as gardening was used as a method of keeping active and occupied. Gardens also provided a space to exercise, which was particularly helpful for those who were clinically vulnerable:

> *“we’ve got two acres of garden…you have to put in a fair amount of work…the garden is really good for weightlifting and everything else”* (Age 55-59, LTC)

Conversely, lack of garden access served as a barrier to PA:

> *“Exercising has definitely been a problem, because of not being able to go out and being stuck in a small flat.”* (Age 35-39, LTC)

##### 1.2 Living in Rural vs Urban Environments

PA was easier for those living in rural locations, where activities such as walking were viewed as both exercise and an enjoyable activity:

> *“We live in a semi-rural (location) and we’ve got beautiful walks near us that are five minutes away. So, I’m exercising every day”* (Age 65-69, LTC)

Rural environments were also conducive to social distancing and enabled participants to feel safe engaging in PA:

> *“we’ve got a lot more green space. So, when I want to go for a jog I can still explore for ages and not see another human being. So, it feels safe…”* (Age 25-29, MHC).

Spending time in nature and gratitude for the natural environment was a comfort to some participants during a time of restriction and this encouraged them to go outside:

> “*I want to get out and get fresh air, you can see the season changing*.” (Age 70-74, Older Adult)

In contrast, living in urban environments was a barrier to PA and provided less physical opportunity due to a lack of open public spaces, limited garden access and crowded environments that were not conducive to social distancing:

> *“They’re all terraced houses here. Everyone lives on top of each other. It just isn’t possible to go out and try and exercise while socially distancing.”* (Age 30-34, MHC)

##### 1.3 Influence of Weather

Warm and sunny weather during the first UK COVID-19 lockdown facilitated PA as activities such as walking were experienced as more pleasant:

> *“I can go for walks with friends*…*I haven’t taken advantage of that a lot recently, but I did when lockdown first came up, because the weather was nicer.”* (Age 45-49, Parent)

Others felt that the hot weather made exercise more difficult. One participant also reflected on how poor weather meant they reduced their PA:

> *“for the bit of March and April, and most of May, I was going out once every two days, sometimes if the weather wasn’t good, once every three days”*. (Age 65-69, LTC)

#### 2. Impact of COVID-19 restrictions

COVID-19 restrictions were a substantial barrier to PA for many participants. However, those with LTCs described being particularly impacted by the shielding advice and guidance to follow social distancing rules more stringently. Conversely, for some participants, restrictions provided a reason to go outside, which was particularly important for parents of young children who were working from home.

##### 2.1 Government Guidance for Clinically Vulnerable Populations (Shielding Advice)

For some participants with a LTC and older adults, the ‘shielding’ advice prompted them to stay inside their homes for extensive periods of time and some did not even use their gardens due to a literal interpretation of guidance:

> *“I got all the texts from the government telling me that I shouldn’t leave the house. And I shouldn’t even put the bins out, or go in the garden if anyone else was going to be around. And so I did follow that really strictly. I didn’t even go in the garden for about six weeks”* (Age 30-34, LTC)

Therefore, for clinically vulnerable populations, environmental facilitators such as garden access may not have been enough to offset the barrier of shielding for PA due to lack of clear guidance.

Shielding also restricted PA for participants’ family members:

> *“My mum’s been like, can I go for a walk? No, unfortunately, you can’t. If I can’t go for a walk, you have to live the same life as me, which means you can’t go for a walk.”* (Age 35-39, LTC)

##### 2.2 Restrictions Around Exercise

For those participants who regularly engaged in team sports and local groups before the pandemic, the closure of group sports activities was described as a barrier to PA:

> *“The one thing that’s been really affected lockdown for me was athletics. I do 400 metres training, so I would run at the track in the evenings. But the UK Athletics when social distancing began, put in decisions to not allow sessions to be run in person anymore.”* (Age 20-24, Young Adult)

Legal restrictions during the pandemic also included enforced closure of sports facilities. Several participants spoke about how they previously used gyms but were unable to as gyms closed:

> *“So obviously my gym closed down so I didn’t do any of that.”* (Age 18-19, Young Adult)

Therefore, participants had a reduced opportunity to engage in PA as their usual resources for exercising were removed. Some participants also shared a feeling of confusion towards the national restrictions and felt that the fear of being punished for not following guidance impacted their enjoyment of PA:

> *“I drove to a park a couple of times and ran around there, but I think there was such public controversy about people driving to other areas that it just made it really not enjoyable”*. (Age 30-34, MHC)
>
> *“I should be playing tennis now…you can only play singles unless you’ve got four in the same household…You rely on somebody else then, to find somebody else to play singles with you, when you’re on your own…And I haven’t found anybody yet*…*So, confusing, shall we say?”* (Age 70-74, Older Adult)

##### 2.3 Government Guidance to Work from Home

For some participants, pandemic restrictions or shielding guidance for people with LTCs meant that they had to work from home. This was referred to as a barrier to PA by each group except older adults who were mostly retired. Some participants felt that working from home also led to less incidental PA:

> *“I had started to gain weight at the beginning of the lockdown, and I realised the very little exercise that I was doing before was the walking to my (university) classes or walking to travel to my classes”* (Age 20-24, Young Adult)

Therefore, as participants had a reduced necessity to engage in PA during the pandemic, this contributed to reduced activity levels. However, in some cases, working from home led to having more time for PA due to less work-related travel and commuting:

> *“[it’s]…also about having more time. Because my partner would have been sat in an office four days a week nine to five, I would have been doing bits of travelling for my work, sometimes leaving early, coming home late.”* (Age 35-39, Parent)

##### 2.4 A Reason to Leave the Home

Some people described engaging in PA to alleviate boredom at home and viewed exercise as a form of escapism from the ‘stay at home’ order:

> *“I guess the exercise was just as much for getting out as it was for exercise.”* (Age 30-34, Parent)
>
> *“we’ve all made sure we do something outside of our four walls, even if it’s a school run, it gets you out of the house, gets you walking”* (Age 40-44, Parent)

Therefore, for individuals who were not motivated to engage in PA for health and fitness, motivation came from wanting to be outside of their homes and to spend time alone:

> *“I remember the first time I just went for a run on my own, and I was like, oh, it’s just so nice, 40 minutes”* (Age 35-39, Parent)

Furthermore, for some participants, restrictive guidance to exercise once per day actually led to increased PA:

> *“It was easy before to say, I’ll go for a walk tomorrow, whereas now it’s a bit like the forbidden fruit, isn’t it? You’re not allowed out except for one walk so you make sure you get out for that walk”*. (Age 70-74, Older Adult)

#### 3. Fear of contracting COVID-19

Fear of contracting COVID-19 and transmitting the virus to others was described as a barrier to PA, particularly for those living with LTCs and MHCs:

> *“It worries me a lot, other people getting ill as a result, maybe if I got it, I could carry it, transmit it to other people. So, I thought no, I will stay at home…I won’t go outside whatsoever.”* (Age 20-24, MHC/Young Adult)

Conversely, some participants were motivated to keep fit due to fear of COVID-19 related health issues. For example, one individual increased their PA to improve their health specifically in response to the pandemic:

> *“obviously the pandemic is a health thing. It just made me realise how important it is to look after yourself”*. (Age 25-29, MHC)

This feeling was heightened for those with increased health risk from COVID-19 due to pre-existing LTCs:

> *“when the news came out about the greater risks (of COVID-19) for diabetes, I’ve switched to going (walking) every day, unless I absolutely can’t”*. (Age 65-69, LTC)

#### 4. Level of engagement with home exercise

Closures of sports facilities and cancellation of group sports meant that some participants turned to alternative means of exercise by attending online exercise classes and purchasing exercise equipment. This was mentioned by all groups but was particularly important for people with LTCs.

Some individuals made significant use of online exercise resources during the pandemic:

> *“Obviously we took the classes online, which has been fantastic*…*It’s as wonderful as if we were all in the class together, it’s been great.”* (Age 65-69, LTC)

Therefore, greater awareness, access and use of online PA resources during the pandemic may have provided an opportunity to engage in new types of PA. Additionally, for one participant, the online classes were easier to access:

> *“Pilates was run at my office, but that was full. So, I couldn’t get a place, but when it went online, she had more capacity, so I could actually do it.”* (Age 45-49, Parent)

Furthermore, some participants purchased exercise equipment which facilitated PA engagement when exercise facilities closed:

> *“When we realised that lockdown was coming, we went and bought the Pilates machine because we used to go to a weekly one to one Pilates class locally.”* (Age 60-64, LTC)

However, making use of virtual classes and home exercise equipment was not a viable option for all participants:

> *“I did look into getting an exercise bike. It was delivered and…downstairs came upstairs to complain about that.”* (Age 40-44, MHC)
>
> *“My daughter was supposed to do a virtual dancing, but I ruled it out straight away because of the fact that we’re in such a small property, with so many of us.”* (Age 30-34, Parent/MHC)

Therefore, living circumstances prohibited some individuals from using home exercise equipment and attending virtual classes. Some also felt that virtual classes failed to replicate the in-person experience, which affected their engagement:

> *“I tried to do Tai-Chi with my teacher, an online link and video. But it just is not the same. So, I stopped doing that.”* (Age 35-39, Parent)

#### 5. Caring responsibilities and conflicting priorities

Parents of young children had increased caring responsibilities during the pandemic due to school closures and lack of childcare, and mentioned this as a barrier to PA:

> *“So, previously, I could take my daughter to a breakfast club very regularly, and that would allow me to go to the gym before work…therefore it started to become very difficult without a breakfast club.”* (Age 45-49, Parent)

For some, having greater demands on their daily lives due to lack of childcare meant that PA was deemed less important than other tasks:

> *“I used to run a lot*…*Since not having an au pair it’s been more difficult. I started running again but it’s just having the time. There’s something’s got to give in a day and it’s usually not having time to do the garden or not going for a run if I’ve got things to do.”* (Age 45-49, Parent/LTC)

Furthermore, some felt that working from home led to them spending more of their free time catching up on work tasks which reduced time for PA:

> *“there has been a difficulty of separating work from home*…*You keep thinking, just one more email that I really need to respond to and that just snowballs. You just don’t know when to stop.”* (Age 40-44, LTC)

#### 6. PA as a method of socialising

For some participants, the inability to continue with usual ways of socialising (e.g. visiting people’s homes and hospitality venues) during the pandemic led to using walking with others as a way of socialising. Parents and younger adults described using PA as a reason to socialise with others:

> *“I was going to circuit training once, two days a week*…*It was more for the social side of it rather than fitness”* (Age 20-24, Young Adult)
>
> *“I’ve got a friend who just lives across the railway tracks and we meet up quite frequently because it’s just a case of just walking up the road and we can meet at the canal and go for a walk.”* (Age 30-34, Parent)

For some participants, using walking to socialise led to increased PA:

> *“Walking has become a massive thing…I try to go for a walk at least every day, I’ve never done that before. So, we’ve found other ways that we can socially interact or meet up with each other, rather than just sitting in the pub”*. (Age 40-44, MHC)

In addition, moves to online delivery of exercise classes and clubs meant that participants were able to socialise while exercising:

> *“I’ve done the Keep Fit, and other people from work do it as well, so we see each other on Zoom and we laugh about it.”* (Age 50-54, LTC)

#### 7. Establishing new routines

The use of routine as a facilitator for PA was mentioned across all groups except young adults. Several individuals reported making walking a part of their daily routine during the pandemic. One participant shared:

> *“…actually getting out for exercise on a daily basis becomes more of a part of what we do.”* (Age 70-74, LTC)

This may have also improved participants’ health and fitness:

> *“And I took up the walking locally. I would just leave my house, walk up the road, round the block and come back for 20 minutes initially, and then gradually increased that to the point where I’m now doing up to an hour.”* (Age 80-84, Older Adult)

Furthermore, those who established new habits and increased their PA suggested that they may be more likely to exercise as restrictions eased:

> *“I wonder if this is something that we will take post-pandemic because now having exercised every day feels like it’s an absolute must have.”* (Age 35-39, Parent)

#### 8. The importance of PA for protecting mental health

Many participants felt that PA had a positive effect on their mental health during the pandemic. This was one of the most important facilitators for most of the demographic groups, apart from older adults who referred to this less frequently.

Some felt that the positive effect of PA on mental health was particularly relevant when having to work from home, as walking gave a structure to their day:

> “*I did go out for the once a day exercise. I usually tried to do it after work just to give me that break to change from my mindset from work mode to coming back to relaxing”*. (Age 20-24, Young Adult)

Furthermore, some participants also described feeling overwhelmed by home confinement which was managed by PA:

> *“I think that’s why my mental health’s been kind of okay after those few weeks when we started going out for a walk, because it was too much to just be totally indoors”* (Age 30-34, LTC)

## Discussion

This study aimed to explore barriers and facilitators to PA among different demographic groups whose PA was likely affected by the COVID-19 pandemic, based on previous research and pandemic restrictions. The study was conducted to give context to existing quantitative research and aimed to map identified barriers and facilitators onto the COM-B model to inform intervention development for protecting PA levels in the current pandemic environment, and during future lockdowns and pandemics.

Most themes primarily mapped onto the physical opportunity and reflective motivation COM-B domains. No themes related to capability for PA. These findings are consistent with a survey study on determinants of PA during the COVID-19 pandemic, which found that physical opportunity and reflective motivation were the most consistent predictors of behaviour (Spence et al., 2021). Knight et al (2021) also found that many themes related to physical opportunity. However, they also found that several themes mapped onto physical or psychological capability. The difference in findings for the present study may be explained by the fact that Knight et al (2021) looked at secondary data from quantitative studies, meaning an understanding of what underpinned apparent relationships was limited. For example, the reported association between mental health difficulties and reduced PA may be explained by fear of contracting COVID-19 (automatic and reflective motivation), rather than a lack of psychological capability.

In the present study, a key theme was the importance of the outdoor environment. Garden access was helpful for PA, with lack of garden access hindering PA, which aligns with previous research findings (Smith et al., 2021). Furthermore, those living in rural areas reported these surroundings were conducive to PA engagement, compared with urban areas with greater population density and higher COVID-19 infection rates (CDC COVID-19 Response Team, 2020; Public Health England, 2020; Hughes & Hughes, 2020; Knight et al., 2021; Goethals et al., 2020). However, the present study builds on previous findings as insights were provided from those who felt that living in/near rural locations benefited their PA levels as they were able to follow social distancing guidance. Therefore, it may be helpful for future pandemic guidance to encourage the use of gardens and local open spaces as helpful environments for PA. Additionally, future guidance could differ regionally so that those living in urban areas without garden access are allowed to travel further to exercise safely. Alternatively, the UK may also benefit from adopting some PA promotion initiatives such as video guidance for exercising at home and outside under pandemic restrictions, as implemented by other European countries (European Network for the Promotion of Health-Enhancing Physical Activity, 2020).

Participants also felt that COVID-19 restrictions were an important factor for explaining changes in their PA levels, which relates to prior research findings (Greenwood-Hickman et al., 2021; Ng et al., 2020; Ng et al., 2021; Smith et al., 2021; Knight et al., 2021). However, the current study develops these findings by highlighting that those with LTCs were particularly impacted by restrictions due to shielding guidance. In future pandemics, it may be useful to advise vulnerable groups on how they can maintain their PA in a safe way (e.g., exercising within the home and in gardens, using virtual resources or home exercise equipment etc.). Furthermore, an individual in our study who was advised to ‘shield’ reported not even going into their garden which suggests that clearer guidance around the relative safety of using well-ventilated areas for PA is key. Furthermore, this study showed that PA as a reason to leave the home was particularly key for parents of young children who were working from home as it provided an opportunity to be outside and spend time alone. Therefore, to engage parents who struggled to make time for PA during the pandemic due to increased caring responsibilities, future interventions may benefit from offering incentives for those who commit to engaging in PA, in line with the Vitality Active Rewards intervention (Hajat, Hasan, Subel & Noach, 2019), or from providing further guidance on how to exercise with children.

The importance of PA for protecting mental health was a common facilitator in this study, which has also been identified by previous research (Greenwood-Hickman et al., 2021; Ng et al., 2021). However, our study also found that older adults referred to this less than the other groups. This could be explained by the pandemic potentially having a less significant effect on their mental health, or perhaps older adults were better able to cope with the effects of the pandemic (González-Sanguino et al., 2020; Klaiber, Wen, DeLongis & Sin, 2021). Alternatively, this could be because mental health awareness and literacy is typically lower in these groups (Malkin et al., 2019). Education on the effect of the pandemic on mental health, and the benefits of regular PA on mental health may be a helpful way to encourage at-risk populations to be more active. This is particularly relevant as rates of mental health difficulties in the UK increased substantially during periods of heightened restrictions (Kwong et al., 2020; O’Connor et al., 2020; Office for National Statistics, 2021). Therefore, consideration for how PA benefits MH during pandemics (Zhang, Zhang, Ma & Di, 2020; Coyle, Ghazi & Georgiou, 2020) may prevent additional burden on health services.

In addition, fear of contracting COVID-19 was an established barrier to PA (Goethals et al., 2020; Smith et al., 2021). However, our study demonstrated that this was a barrier particularly for those with LTCs and MHCs, who may subsequently benefit from targeted support around exercising at home. The current study also showed a new perspective: that some people with LTCs increased their PA due to the increased focus on public health and awareness around the effects of COVID-19 on LTCs. Therefore, modelling using examples of those with LTCs who increased their PA during the pandemic may be an effective way of encouraging other individuals with LTCs (Michie et al., 2011).

Finally, it is important to consider the merits and limitations of this study. Particularly, this study had a large sample size of 116 semi-structured interviews, which enabled inclusion of a broad range of views and experiences. Furthermore, three researchers met regularly throughout data analysis to discuss themes and to ensure that thematic analysis was applied appropriately. Thirdly, this study gives context to the quantitative findings relating to PA during the COVID-19 pandemic. Fourthly, in conjunction with the COM-B model (Michie et al., 2011), this study can be used to consider intervention components for increasing PA in future lockdowns and pandemics. However, this study also has several limitations. Firstly, the interviews were with individuals who were predominantly female, white British and educated to degree level, which limits the range of perspectives represented. The use of online and telephone interviews may also have excluded those who did not have online access. Additionally, as the CSS focused on broader mental health effects of the pandemic on several aspects of life, the depth of reflections relating to PA may be limited, however all participants were explicitly asked to reflect on their PA both before and during the pandemic.

In conclusion, this study is the first to explore barriers and facilitators to PA for different demographic groups during the COVID-19 pandemic, and to map these onto a theoretical framework to aid understanding of PA behaviours and to inform future interventions. Future interventions should aim to harness the facilitating factors and off-set barriers to PA with strategies including improving access to home exercise resources, tailoring PA guidance for areas with higher population density and providing education on the positive physical and mental effects of PA. This could reduce further long-term health issues resulting from reduced PA and may be a protective factor against worsening mental health during times of social isolation.

## Data Availability

The data are not publicly available due to their containing information that could compromise
the privacy of research participants

## Acknowledgements

The authors would like to thank the participants from the COVID-19 Social Study and the organisations who supported recruitment including; the MARCH Mental Health Research Network, the TRIUMPH network, MQ Participate, the National Elf Service, Age Concern, The Healthy Aging Initiative, The Baring Foundation and the carers’ research volunteer network of the Alzheimer’s Society. The researchers are also grateful to Alison McKinlay, Anna Roberts, Tom May, Jo Dawes, Sara Esser, Rana Conway and Henry Aughterson who conducted the COVID-19 Social Study interviews.

## References

Assaloni, R., Pellino, V. C., Puci, M. V., Ferraro, O. E., Lovecchio, N., Girelli, A. & Vandoni, M. (2020). Coronavirus disease (Covid-19): How does the exercise practice in active people with type 1 diabetes change? A preliminary survey. Diabetes research and clinical practice, 166, 108297. https://doi.org/10.1016/j.diabres.2020.108297

Atkins, L., Francis, J., Islam, R., O’Connor, D., Patey, A., Ivers, N., Foy, R., Duncan, E. M., Colquhoun, H., Grimshaw, J. M., Lawton, R. & Michie, S. (2017). A guide to using the Theoretical Domains Framework of behaviour change to investigate implementation problems. Implementation Science, 12(1), 77. https://doi.org/10.1186/s13012-017-0605-9

Braun, V. & Clarke, V. (2006). Using thematic analysis in psychology. Qualitative Research in Psychology, 3(2), 77–101. https://doi.org/10.1191/1478088706qp063oa

Braun, V. & Clarke, V. (2019). Reflecting on reflexive thematic analysis. Qualitative Research in Sport, Exercise and Health, 11(4), 589–597. https://doi.org/10.1080/2159676X.2019.1628806

Braun, V. & Clarke, V. (2021). One size fits all? What counts as quality practice in (reflexive) thematic analysis? Qualitative Research in Psychology, 18(3), 328–352. https://doi.org/10.1080/14780887.2020.1769238

Burton, A., McKinlay, A., Aughterson, H. & Fancourt, D. (2021). Impact of the COVID-19 pandemic on the mental health and well-being of adults with mental health conditions in the UK: a qualitative interview study. Journal of Mental Health, 1–8. https://doi.org/10.1080/09638237.2021.1952953

Burton, A., McKinlay, A., Dawes, J., Roberts, A., Fynn, W., May, T. & Fancourt, D. (2022). Understanding barriers and facilitators to compliance with UK social distancing guidelines during the COVID-19 pandemic: A qualitative interview study. Behaviour Change, 1–21. doi: 10.1017/bec.2021.27

Carvalho, J., Borges-Machado, F., Pizarro, A. N., Bohn, L. & Barros, D. (2021). Home Confinement in Previously Active Older Adults: A Cross-Sectional Analysis of Physical Fitness and Physical Activity Behavior and Their Relationship With Depressive Symptoms. Frontiers in Psychology, 12, Article 643832. https://doi.org/10.3389/fpsyg.2021.643832

CDC COVID-19 Response Team (2020). Geographic Differences in COVID-19 Cases, Deaths, and Incidence – United States, February 12-April 7, 2020. MMWR. Morbidity and mortality weekly report, 69(15), 465–471. https://doi.org/10.15585/mmwr.mm6915e4

Covid-19 Social Study (2020). Home. Retrieved from https://www.covidsocialstudy.org/

Coyle, C., Ghazi, H. & Georgiou, I. (2020). The mental health and well-being benefits of exercise during the COVID-19 pandemic: a cross-sectional study of medical students and newly qualified doctors in the UK. Irish Journal of Medical Science, 190(3), 925–926. https://doi.org/10.1007/s11845-020-02423-z

Creese, B., Khan, Z., Henley, W., O’Dwyer, S., Corbett, A., Da Silva, M. V., Mills, K., Wright, N., Testad, I., Aarsland, D. & Ballard, C. (2021). Loneliness, physical activity, and mental health during COVID-19: a longitudinal analysis of depression and anxiety in adults over the age of 50 between 2015 and 2020. International Psychogeriatrics, 33(5), 505–514. https://doi.org/10.1017/S1041610220004135

Curtis, R. G., Olds, T., Ferguson, T., Fraysse, F., Dumuld, D., Esterman, A., Hendrie, G. A., Brown, W. J., Lagiseti, R. & Maher, C. A. (2021). Changes in diet, activity, weight and wellbeing of parents during COVID-19 lockdown. PloS ONE, 16(3), e0248008. https://doi.org/10.1371/journal.pone.0248008

Dawes, J., May, T., McKinlay, A., Fancourt, D. & Burton, A. (2021). Impact of the COVID-19 pandemic on the mental health and wellbeing of parents with young children: a qualitative interview study. BMC Psychology, 9(1), 194. https://doi.org/10.1186/s40359-021-00701-8

European Network for the Promotion of Health-Enhancing Physical Activity (2020). HEPA Europe Newsletter May 2020. Retrieved from https://www.euro.who.int/en/health-topics/disease-prevention/physical-activity/activities/hepa-europe/subscribe-to-receive-our-news-alerts

Fisher, A., Roberts, A., McKinlay, A., Fancourt, D. & Burton, A. (2021). The impact of the COVID-19 pandemic mental health and well-being of people living with a long-term physical health condition: a qualitative study. BMC Public Health, 21(1), 1801. https://doi.org/10.1186/s12889-021-11751-3

Ghosh, A., Arora, B., Gupta, R., Anoop, S. & Misra, A. (2020). Effects of a nationwide lockdown during COVID-19 epidemic on lifestyle and other medical issues of patients with type 2 diabetes in north India. Diabetes & metabolic syndrome, 14(5), 917–920. https://doi.org/10.1016/j.dsx.2020.05.044

Goethals, L., Barth, N., Guyot, J., Hupin, D., Celarier, T. & Bongue, B. (2020). Impact of Home Quarantine on Physical Activity Among Older Adults Living at Home During the COVID-19 Pandemic: Qualitative Interview Study. JMIR aging, 3(1), e19007. https://doi.org/10.2196/19007

González-Sanguino, C., Ausín, B., Castellanos, M.Á., Saiz, J., López-Gómez, A., Ugidos, C. & Muñoz, M. (2020). Mental health consequences during the initial stage of the 2020 Coronavirus pandemic (COVID-19) in Spain. Brain, behavior and immunity, 87, 172–176. https://doi.org/10.1016/j.bbi.2020.05.040

GOV UK (2021). Prime Minister announces national lockdown. Retrieved from https://www.gov.uk/government/news/prime-minister-announces-national-lockdown

Greenwood-Hickman, M. A., Dahlquist, J., Cooper, J., Holden, E., McClure, J. B., Mettert, K. D., Perry, S. R. & Rosenberg, D. E. (2021). “They’re Going to Zoom It”: A Qualitative Investigation of Impacts and Coping Strategies During the COVID-19 Pandemic Among Older Adults. Frontiers in Public Health, 9, 679976. https://doi.org/10.3389/fpubh.2021.679976

Hajat, C., Hasan, A., Subel, S. & Noach, A. (2019). The impact of short-term incentives on physical activity in a UK behavioural incentives programme. npj Digital Medicine, 2(1), Article 91. https://doi.org/10.1038/s41746-019-0164-3

Howlett, N., Schulz, J., Trivedi, D., Troop, N. & Chater, A. (2017). A prospective study exploring the construct and predictive validity of the COM-B model for physical activity. Journal of Health Psychology, 24(10), 1378–1391. https://doi.org/10.1177/1359105317739098

Hughes, R. P. & Hughes, D. A. (2020). Impact of Relaxing Covid-19 Social Distancing Measures on Rural North Wales: A Simulation Analysis. Frontiers in Public Health, 8, Article 562473. https://doi.org/10.3389/fpubh.2020.562473

Institute for Government (2021). Schools and coronavirus: The government’s handling of education during the pandemic. Retrieved from https://www.instituteforgovernment.org.uk/sites/default/files/publications/schools-and-coronavirus.pdf

Klaiber, P., Wen, J. H., DeLongis, A. & Sin, N. L. (2021). The Ups and Downs of Daily Life During COVID-19: Age Differences in Affect, Stress, and Positive Events. The journals of gerontology, Series B, Psychological sciences and social sciences, 76(2), e30–e37. https://doi.org/10.1093/geronb/gbaa096

Knight, R. L., McNarry, M. A., Sheeran, L., Runacres, A. W., Thatcher, R., Shelley, J. & Mackintosh, K. A. (2021). Moving Forward: Understanding Correlates of Physical Activity and Sedentary Behaviour during COVID-19 – An Integrative Review and Socioecological Approach. International journal of environmental research and public health, 18(20), 10910. https://doi.org/10.3390/ijerph182010910

Kwong, A. S. F., Pearson, R. M., Adams, M. J., Northstone, K., Tilling, K., Smith, D., Fawns-Ritchie, C., Bould, H., Warne, N., Zammit, S., Gunnell, D. J., Moran, P. A., Micali, N., Reichenberg, A., Hickman, M., Rai, D., Haworth, S., Campbell, A., Altschul, D., Flaig, R., McIntosh, A. M., Lawlor, D. A., Porteous, D. & Timpson, N. J. (2020). Mental health before and during the COVID-19 pandemic in two longitudinal UK population cohorts. The British Journal of Psychiatry, 218(6), 334–343. https://doi.org/10.1192/bjp.2020.242

Malkin, G., Hayat, T., Amichai-Hamburger, Y., Ben-David, B. M., Regev, T. & Nakash, O. (2019). How well do older adults recognise mental illness? A literature review. Psychogeriatrics, 19(5), 491–504. https://doi.org/10.1111/psyg.12427

MARCH Network (2022). Home page: March Legacy. Retrieved from https://www.marchlegacy.org/

Mata, J., Wenz, A., Rettig, T., Reifenscheid, M., Möhring, K., Krieger, U., Friedel, S., Fikel, M., Cornesse, C., Blom, A. G. & Naumann, E. (2021). Health behaviors and mental health during the COVID-19 pandemic: A longitudinal population-based survey in Germany. Social Science & Medicine, 287, 114333. https://doi.org/10.1016/j.socscimed.2021.114333

McCarthy, H., Potts, H. W. W. & Fisher, A. (2021). Physical Activity Before, During and After COVID-19 Restrictions: Longitudinal Smartphone-Tracking Study of Adults in the United Kingdom. Journal of medical internet research, 23(2), Article e23701. https://doi.org/10.2196/23701

McKinlay, A., Fancourt, D. & Burton, A. (2021). A qualitative study about the mental health and wellbeing of older adults in the UK during the COVID-19 pandemic. BMC Geriatrics, 21(1), 439. https://doi.org/10.1186/s12877-021-02367-8

McKinlay, A., May, T., Dawes, J., Fancourt, D. & Burton, A. (2021). “You’re just there, alone in your room with your thoughts” A qualitative study about the impact of lockdown among young people during the COVID-19 pandemic. medRxiv. https://doi.org/10.1101/2021.04.11.21254776

Michie, S., van Stralen, M. M. & West, R. (2011). The behaviour change wheel: A new method for characterising and designing behaviour change interventions. Implementation Science, 6, 42. https://doi.org/10.1186/1748-5908-6-42

Mishra, R., Park, C., York, M. K., Kunik, M. E., Wung, S., Naik, A. D. & Najafi, B. (2021). Decrease in Mobility during the COVID-19 Pandemic and Its Association with Increase in Depression among Older Adults: A Longitudinal Remote Mobility Monitoring Using a Wearable Sensor. Sensors, 21(9), 3090. https://doi.org/10.3390/s21093090

Ng, K., Cooper, J., McHale, F., Clifford, J. & Woods, C. (2020). Barriers and facilitators to changes in adolescent physical activity during COVID-19. BMJ Open Sport & Exercise Medicine, 6, e000919. https://doi.org/10.1136/bmjsem-2020-000919

Ng, Y. L., Hill, K. D. & Burton, E. (2021). Exploring physical activity changes and experiences of older adults living in retirement villages during a pandemic. Australasian journal on ageing. https://doi.org/10.1111/ajag.12963

O’Connor, R. C., Wetherall, K., Cleare, S., McClelland, H., Melson, A. J., Niedzwiedz, C. L., O’Carroll, R. E., O’Connor, D. B., Platt, S., Scowcroft, E., Watson, B., Zortea, T., Ferguson, E. & Robb, K. A. (2020). Mental health and well-being during the COVID-19 pandemic: longitudinal analyses of adults in the UK COVID-19 Mental Health & Wellbeing study. The British Journal of Psychiatry, 218(6), 326–333. https://doi.org/10.1192/bjp.2020.212

Office for National Statistics (2020). Coronavirus and shielding of clinically extremely vulnerable people in England: 28 May to 3 June 2020. https://www.ons.gov.uk/peoplepopulationandcommunity/healthandsocialcare/conditionsanddiseases/bulletins/coronavirusandshieldingofclinicallyextremelyvulnerablepeopleinengland/28mayto3june2020

Office for National Statistics (2021). Coronavirus and depression in adults, Great Britain: January to March 2021. https://www.ons.gov.uk/peoplepopulationandcommunity/wellbeing/articles/coronavirusanddepressioninadultsgreatbritain/januarytomarch2021

Public Health England (2020). Disparities in the risk and outcomes of COVID-19. https://assets.publishing.service.gov.uk/government/uploads/system/uploads/attachment_data/file/908434/Disparities_in_the_risk_and_outcomes_of_COVID_August_2020_update.pdf

Robbins, L. B. & Ling, J. (2022). Lifestyle Behaviors and Parents’ Mental Wellbeing Among Low-Income Families During COVID-19 Pandemic. Nursing Research, 10.1097/NNR.0000000000000576. Advance online publication. https://doi.org/10.1097/NNR.0000000000000576

Roberts-Lewis, S. F., Ashworth, M., White, C. M. & Rose, M. R. (2021). COVID-19 lockdown impact on the physical activity of adults with progressive muscle diseases. BMJ Neurology Open, 3(1), e000140. https://doi.org/10.1136/bmjno-2021-000140

Rogers, N. T., Waterlow, N. R., Brindle, H., Enria, L., Eggo, R. M., Lees, S. & Roberts, C. H. (2020). Behavioral Change Towards Reduced Intensity Physical Activity Is Disproportionately Prevalent Among Adults With Serious Health Issues or Self-Perception of High Risk During the UK COVID-19 Lockdown. Frontiers in public health, 8, 575091. https://doi.org/10.3389/fpubh.2020.575091

Savage, M. J., Hennis, P. J., Magistro, D., Donaldson, J., Healy, J. C. & James, R. M. (2021). Nine Months into the COVID-19 Pandemic: A Longitudinal Study Showing Mental Health and Movement Behaviours Are Impaired in UK Students. International Journal of Environmental Research and Public Health, 18(6), 2930. https://doi.org/10.3390/ijerph18062930

Schmidt, T. & Pawlowski, C. S. (2021). Physical Activity in Crisis: The Impact of COVID-19 on Danes’ Physical Activity Behaviour. Frontiers in sports and active living, 2, 610255. https://doi.org/10.3389/fspor.2020.610255

Smith, L., Jacob, L., Butler, L., Schuch, F., Barnett, Y., Grabovac, I., Veronese, N., Caperchione, C., Lopez-Sanchez, G. F., Meyer, J., Abufaraj, M., Yakkundi, A., Armstrong, N. & Tully, M. A. (2020). Prevalence and correlates of physical activity in a sample of UK adults observing social distancing during the COVID-19 pandemic. BMJ Open Sport & Exercise Medicine, 6, e000850. https://doi.org/10.1136/bmjsem-2020-000850

Smith, T. O., Belderson, P., Dainty, J. R., Birt, L., Durrant, K., Chipping, J. R., Tsigarides, J., Yates, M., Naughton, F., Werry, S., Notley, C., Shepstone, L. & MacGregor, A. J. (2021). Impact of COVID-19 pandemic social restriction measures on people with rheumatic and musculoskeletal diseases in the UK: a mixed-methods study. BMJ Open, 11, e048772. https://doi.org/10.1136/bmjopen-2021-048772

Spence, J. C., Rhodes, R. E., McCurdy, A., Mangan, A., Hopkins, D. & Mummery, W. K. (2021). Determinants of physical activity among adults in the United Kingdom during the COVID-19 pandemic: The DUK-COVID study. British journal of health psychology, 26(2), 588–605. https://doi.org/10.1111/bjhp.12497

Stockwell, S., Trott, M., Tully, M., Shin, J., Barnett, Y., Butler, L., McDermott, D., Schuch, F. & Smith, L. (2020). Changes in physical activity and sedentary behaviours from before to during the COVID-19 pandemic lockdown: a systematic review. BMJ Open Sport & Exercise Medicine, 7, e000960. https://doi.org/10.1136/bmjsem-2020-000960

Strain, T., Sharp, S. J., Spiers, A., Price, H., Williams, C., Fraser, C., Brage, S., Wijndaele, N.K. & Kelly, P. (2022). Population level physical activity before and during the first national COVID-19 lockdown: A nationally representative repeat cross-sectional study of 5 years of Active Lives data in England. The Lancet Regional Health. Europe, 12, 100265. https://doi.org/10.1016/j.lanepe.2021.100265

Sundler, A. J., Lindberg, E., Nilsson, C. & Palmér, L. (2019). Qualitative thematic analysis based on descriptive phenomenology. Nursing Open, 6(3), 733–739. https://doi.org/10.1002/nop2.275

To, Q. G., Duncan, M. J., Van Itallie, A. & Vandelanotte, C. (2021). Impact of COVID-19 on Physical Activity Among 10,000 Steps Members and Engagement With the Program in Australia: Prospective Study. Journal of medical Internet research, 23(1), e23946. https://doi.org/10.2196/23946

Wang, Y., Zhang, Y., Bennell, K., White, D. K., Wei, J., Wu, Z., He, H., Liu, S., Luo, X., Hu, S., Zeng, C. & Lei, G. (2020). Physical Distancing Measures and Walking Activity in Middle-aged and Older Residents in Changsha, China, during the COVID-19 Epidemic Period: Longitudinal Observational Study. Journal of medical Internet research, 22(10), e21632. https://doi.org/10.2196/21632

World Health Organisation (2021). WHO Coronavirus (COVID-19) Dashboard. https://covid19.who.int/

Zhang, Y., Zhang, H., Ma, X. & Di, Q. (2020). Mental Health Problems during the COVID-19 Pandemics and the Mitigation Effects of Exercise: A Longitudinal Study of College Students in China. International Journal of Environmental Research and Public Health, 17(10), 3722. https://doi.org/10.3390/ijerph17103722

